# Co-occurrence of apathy and impulsivity in progressive supranuclear palsy

**DOI:** 10.1101/2021.02.26.21252350

**Authors:** Zi Qi Kok, Alexander G. Murley, Timothy Rittman, James B. Rowe, Luca Passamonti

## Abstract

**Background:** Apathy and impulsivity are common problems in progressive supranuclear palsy (PSP) and can worsen its prognosis. They can co-exist in the same patients although their concomitant prevalence remains unclear. Their relationship to emotional lability is also unknown.

**Objectives:** To estimate the co-occurrence of apathy and impulsivity and their relationship to emotional lability in PSP. To characterize the demographic, clinical, and cognitive features of PSP patients with apathy and impulsivity.

**Methods:** In a retrospective study of a long-term clinical cohort, we assessed the prevalence of apathy, impulsivity, and emotional lability from clinical interviews, medical records, and contemporary carer questionnaires. N=154 patients with a diagnosis of probable or possible PSP (according to the 2017 Movement Disorder Society criteria) were identified. N=64 of these patients had neuropathological confirmation of PSP. PSP patients with both apathy and impulsivity were compared in terms of demographic, clinical, and cognitive characteristics to PSP patients with either one or neither of these neuropsychiatric features.

**Results:** Apathy and impulsivity co-existed in two-thirds of people with PSP. A fifth displayed emotional lability in addition to apathy and impulsivity. Apathy and impulsivity were more commonly co-expressed than by chance. There was no single demographic, clinical or cognitive feature that distinguished between PSP patients with *versus* patients without apathy and impulsivity.

**Conclusions:** The co-existence of apathy and impulsivity in PSP suggests that these neuropsychiatric features may share similar risk factors and etio-pathogenetic mechanisms. Apathy and impulsivity should be jointly assessed when planning symptomatic treatments for behavioural problems in PSP.

## Introduction

Progressive supranuclear palsy (PSP) is a neurodegenerative disorder caused by neuronal and glial aggregation of hyperphosphorylated 4-R tau isoforms, as part of the spectrum of diseases caused by frontotemporal lobar degeneration^1^. PSP is characterised by postural instability, akinetic-rigidity, and oculomotor dysfunction^1^. Patients with PSP often display prominent neuropsychiatric problems, including apathy and impulsivity^2–4^. In PSP and related disorders, apathy and impulsivity reduce survival^5^ and predict loss of functional independence^6^.

Apathy and impulsivity are common, multifactorial, and commonly co-existent symptoms. For example, apathy has affective, cognitive, and behavioural components^7,8^, with a resultant loss of interest in activities and difficulty initiating actions^9^. Impulsivity encompasses actions that are premature, without foresight of deleterious consequences or made as a result of a failure to inhibit contextually inappropriate responses^10,11^. Apathy and impulsivity tend to occur together, including in PSP^4,12^, frontotemporal dementia^11^, and Parkinson’s disease^13^.

This co-existence of apathy and impulsivity argues against the motivational spectrum hypothesis, with apathy and impulsivity at opposite ends of the spectrum^14,15^. There are several and not necessarily mutually exclusive explanations for their reported co-existence. First, it may be chance, given that both are common.

Second, they may have a shared neuroanatomical basis^12,16^, including the degeneration of analogous limbic fronto-striatal circuits, which represent actions, rewards, and inhibitory control^11^. Third, there may be shared neurochemical systems mediating apathy and impulsivity^11,17^, which would be especially relevant for pharmacological treatments. In clinical practice, dopaminergic agents do not clearly ameliorate apathy or impulsivity^11^, but serotonergic and noradrenergic strategies might be effective^18–20^. Fourth, individuals may express apathy and impulsivity at different times, perhaps alternating between them. Detailed temporal studies are lacking, but if the few actions made by an apathetic individual were premature, high-risk, and contextually disinhibited, this would be best interpreted as truly concurrent apathy and impulsivity.

Despite many studies of apathy and impulsivity in PSP^2,4,12,16,21–24^, basic questions about their co-existence remain unanswered. For example, is their co-existence more frequent than by chance? To what extent do these behavioural problems relate to other contextually inappropriate behaviours, such as emotional lability? Emotional lability refers to sudden, rapid, exaggerated, and uncontrollable bursts of laughing or crying that are spontaneous and often inappropriate to the social context. During these episodes, patients may report congruent emotional feelings (i.e., feeling sad while crying) or an incongruity between the external emotional appearance and their ‘internal’ sentiments (i.e. not feeling sad while crying)^25^. Emotional lability has been linked to lesions in the pontine nuclei, cerebellum, and frontal lobes^26^, but a link to impulsivity has not been established.

This study has three aims. First, to test the hypothesis that apathy and impulsivity co-exist in PSP, at the individual level. Second, to test the association between presence of emotional lability and apathy or impulsivity. Third, to compare demographic, clinical, and cognitive features across PSP patients with or without apathy and impulsivity.

## Methods

### Participants

This study included a sample of n=154 patients with a clinical diagnosis of probable or possible PSP, identified by retrospective re-diagnosis according to the 2017 Movement Disorder Society (MDS) criteria^27^. The data were extracted from research records and electronic medical records at a tertiary referral center, from the Pick’s Disease and Progressive Supranuclear Palsy Prevalence and Incidence protocol (12/EE/0475) and the “Diagnosis and prognosis in Progressive Supranuclear Palsy and Corticobasal Degeneration” protocol (07/Q0102/3).

Sixty-four patients had pathological confirmation of PSP, via the Cambridge Brain Bank. Two patients with a clinical diagnosis of PSP in life were excluded as post-mortem examination showed an alternative diagnosis (respectively, corticobasal syndrome and argyrophilic grain disease). Exclusion criteria were the following: diagnosis of other neurodegenerative conditions including corticobasal syndrome (CBS) (although the PSP-CBS subtype as per the 2017 MDS criteria was included), Lewy body dementia, idiopathic Parkinson’s disease, Alzheimer’s disease, stroke, cancer, and normal pressure hydrocephalus.

Clinical and demographic features included gender, age at symptom onset, disease duration, and PSP subtype^27^. Disease severity, motor symptoms, cognition, and behavioural problems were respectively assessed via the Progressive Supranuclear Palsy-Rating Scale (PSPRS), Unified Parkinson’s Disease Rating Scale (UPDRS-part III), Addenbrooke’s Cognitive Examination-Revised (ACE-R), Frontal Assessment Battery (FAB), and Cambridge Behavioural Inventory-revised (CBI-R). Behavioural changes were also identified throughout qualitative analysis of clinical letters and notes, coded in binary terms (absent or present) based on descriptions of apathy, impulsivity, and emotional lability, as reported in *Appendix 1*. The assessment of behavioural changes was supplemented by scores in the CBI-R (questions on *Motivation* and *Abnormal behaviour)* and PSPRS (questions on withdrawal, irritability, and emotional lability, see *Appendix 3* for details).

We used the ACE-R and FAB scores recorded within 3 months of the first clinical record of apathetic or impulsive behavioural changes. PSP patients without this information recorded within three months of their first behavioural presentation were excluded (n=35). For patients who experienced neither behavioural changes, we employed the scores measured within 3 months of their diagnosis of PSP. People who were unable to complete the assessments, or those lacking PSPRS/UPDRS/ACE-R/FAB measures were also excluded (n=16 patients).

### Statistical analyses

Statistical analyses used IBM SPSS (version 27.0) for frequentist analyses and JASP (Version 0.14) for Bayesian analyses. Chi-squared tests for the co-occurrence of apathy, impulsivity, and emotional lability were conducted. One-way analyses of variance (ANOVAs) were used to compare age at onset, disease duration PSPRS, UPDRS, ACE-R, and FAB scores (at the first presentation) between groups with both apathy and impulsivity, only apathy or impulsivity or neither of them. Bayesian ANOVAs were used to test the relative evidence of null (no difference) and alternate (difference) hypotheses.

## Results

PSP patients comprised six subgroups, as reported in **Supplementary Table 1**. Most participants (74%) had PSP-Richardson’s syndrome. The remainder spanned PSP with CBS features (10%), PSP with frontal presentation (8%), PSP with primary gait freezing (4%), PSP-parkinsonism (2%), or PSP with speech and language presentation (1%).

The prevalence of apathy, impulsivity, and emotional lability is shown in **Figure 1**. Overall, 75% of the patient group (n=116) had impulsive behavioural changes; 79% (n=121) had apathetic behavioural problems; and 29% of patients (n=44) manifested emotional lability. Twenty percent displayed all three behavioural changes. Two-thirds had both apathetic and impulsive behaviour reported.

**Figure 1:**
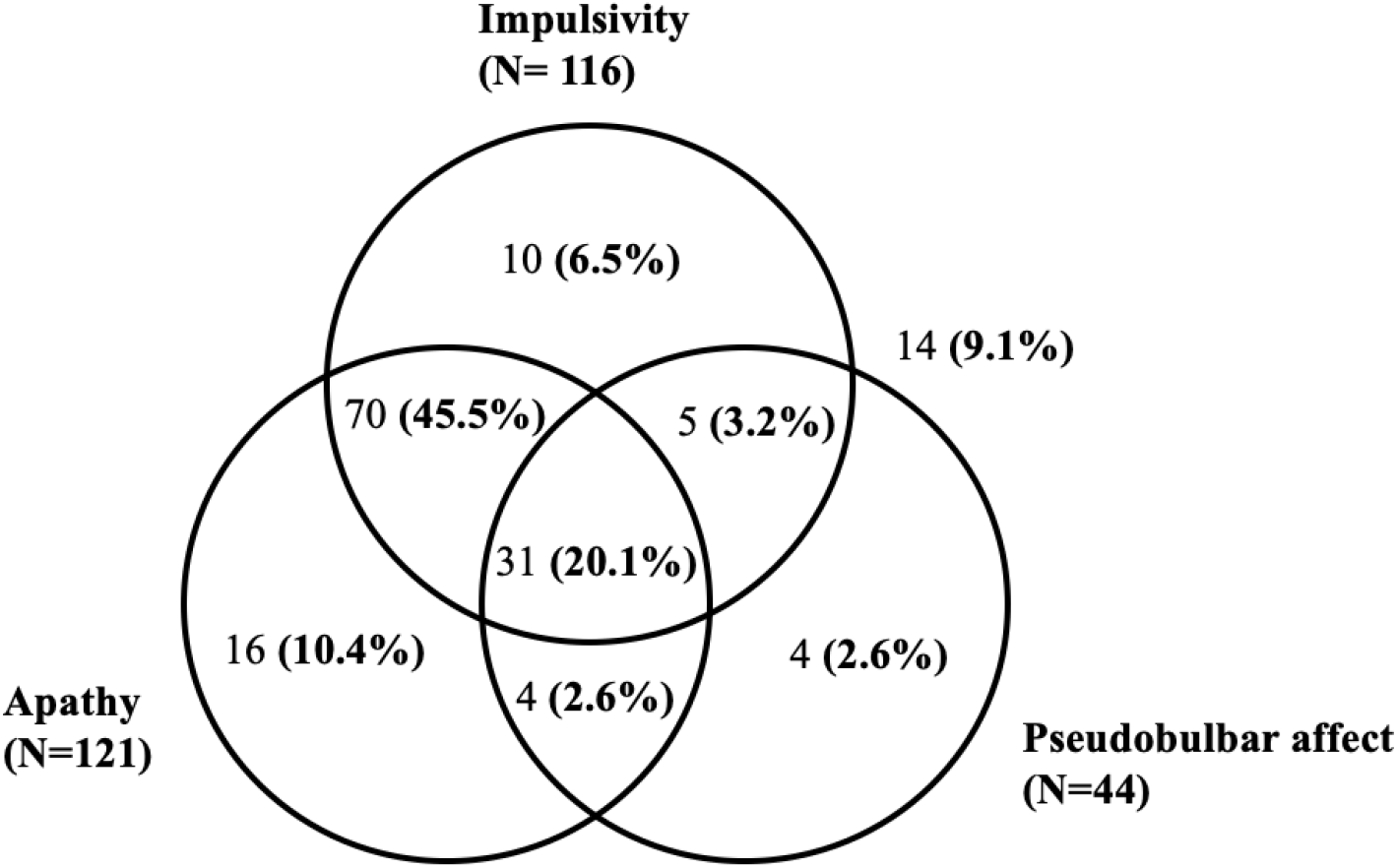
Distribution of patients with apathy, impulsivity, and emotional lability (number of patients given, followed by percentage of total patient sample in parentheses).

**Table 1(A)** shows the relationship between apathy and impulsivity. There was a positive association between them (*χ*^*2*^_*(1,154)*_ *=18*.*2, p*<*0*.*001 continuity corrected)*). In other words, the probability of being apathetic increases with impulsivity. **Table 1(B)** shows the relationship between emotional lability and impulsivity. There was no significant relationship (*χ*^*2*^_*(1,154)*_ <*1, ns)*. In other words, the probability of being impulsive was independent of emotional lability. **Table 1(C)** shows the relationship between emotional lability and apathy. There was no significant relationship (*χ*^*2*^_*(1,154)*_ *<1, ns)*. In other words, the probability of being apathetic was independent of emotional lability. **Table 1(D)** shows the relationship between emotional lability and co-existent apathy and impulsivity. There was no significant relationship (*χ*^*2*^_*(1,154)*_<*1, ns)*. Comparing groups with apathy, impulsivity, neither or both, the frequentist tests (**Supplementary Table 1**), we found no significant difference by gender, age at onset, disease duration, disease severity (as indexed by the PSPRS), motor severity (UPDRS-part III) or global cognitive screening tests (ACE-R, FAB). Bayesian analyses confirmed these null findings with positive or very strong evidence in favor of the null (BF_10_<1/3 or <1/10 respectively; **Supplementary Information**).

**Table 1:** Contingency tables for apathy, impulsivity, and emotional lability in PSP patients

| **A: Contingency table for apathy and impulsivity. $\chi^2_{(1,154)}=18.2, p<0.001$ (continuity corrected)** |  |  |  |  |
| --- | --- | --- | --- | --- |
|  | **Impulsivity (n)** |  |  | **Total (n)** |
|  |  | **No** | **Yes** |  |
| **Apathy (n)** | **No** | 18 | 15 | 33 |
|  | **Yes** | 20 | 101 | 121 |
| **Total** |  | 38 | 116 | 154 |
| **B: Contingency table for emotional lability and impulsivity. $\chi^2_{(1,154)}<1, ns$ (continuity corrected)** |  |  |  |  |
|  | **Emotional lability (n)** |  |  | **Total (n)** |
|  |  | **No** | **Yes** |  |
| **Impulsivity (n)** | **No** | 30 | 8 | 38 |
|  | **Yes** | 80 | 36 | 116 |
| **Total (n)** |  | 110 | 44 | 154 |
| **C: Contingency table for emotional lability and apathy. $\chi^2_{(1,154)}<1$ (corrected)** |  |  |  |  |
|  | **Emotional lability (n)** |  |  | **Total (n)** |
|  |  | **No** | **Yes** |  |
| **Apathy (n)** | **No** | 24 | 9 | 33 |
|  | **Yes** | 86 | 35 | 121 |
| **Total** |  | 110 | 44 | 154 |
| **D: Contingency table for co-existent apathy, impulsivity, and emotional lability. $\chi^2_{(1,154)}<1, ns$ (corrected).** |  |  |  |  |
|  | **Emotional lability (n)** |  |  | **Total (n)** |
|  |  | **No** | **Yes** |  |
| **Co-existent apathy and impulsivity (n)** | **No** | 41 | 13 | 54 |
|  | **Yes** | 69 | 31 | 100 |
| **Total (n)** |  | 110 | 44 | 154 |

## Discussion

This study confirms the high frequency of reported co-existence of apathy and impulsivity in PSP patients, with a positive relationship between them. Neither apathy nor impulsivity were associated with emotional lability. Approximately three-quarters of people with PSP had apathy, in keeping with previous reports that apathy is the most common neuropsychiatric feature of PSP^2–4^. Most patients with apathy also manifested impulsive behaviours, more than by chance association with the prevalence of impulsivity. The prevalence of impulsivity in our cohort was higher than previously reported (74% versus 32% to 43%^3,4,23^). This disparity may be due to methodological differences across studies: patient self-ratings, clinician judgment of behavioural changes, or carer reports using tools such as the Cambridge Behavioural Inventory, Neuropsychiatric Inventory or Frontal Behavioural Inventory.

The strong concomitance of apathy and impulsivity accords with previous studies of other syndromes associated with frontotemporal lobar degeneration^9,12,22,28^ and Parkinson’s disease^9,^. Such a positive relationship across multiple disorders suggests that these neuropsychiatric features may share similar risk factors and etiopathogenetic mechanisms^22,29,30^. For example, apathy and impulsivity have common correlates in the white-matter tracts connecting the prefrontal cortex, basal ganglia, temporal poles, and brainstem^22^; and grey-matter atrophy across the frontal cortex^12,29^. Despite molecular pathological differences, there is convergence onto similar neural circuits. This signals the need to study apathy and impulsivity together rather than in isolation^11,12^, and highlights the potential for joint therapeutic strategies, rather than dopaminergic antagonism between apathy (and akinesia) and impulsivity^31^. In view of their strong positive association, apathy and impulsivity cannot be simply conceptualized as opposite extremes of a dopamine-dependent spectrum, with apathy arising from a hypodopaminergic state and impulsivity from a hyperdopaminergic state^14,15^. Other neurotransmitters are likely to influence apathy and impulsivity. For example, serotonin is reduced in several neurodegenerative disorders, and serotonergic manipulations through serotonin reuptake inhibition can ameliorate deficits in response inhibition in people with Parkinson’s disease and frontotemporal dementia^18,20^. Apathy and impulsivity may also be attributed to noradrenergic deficits^32^. For example, noradrenergic reuptake inhibition improves response inhibition and restores the function of inhibitory control networks in Parkinson’s disease^19,33^.

Although emotional lability was relatively common in PSP (with a prevalence of 20%), it was not associated with either apathy or impulsivity. This suggests that emotional lability arises from dysfunctions in separate neural systems. For example, whilst fronto-striatal systems are affected in apathy and impulsivity^11^, fronto-ponto-cerebellar circuits have been linked to emotional lability or pseudobulbar affect^34^. Both of these neuroanatomical systems show diffuse tau pathology and neurodegeneration in PSP.

We found no specific demographic or clinical characteristics linked to apathy and impulsivity in PSP. This was unexpected as other studies have found associations between apathy and executive function in PSP^4,35 4^. Although it is difficult to directly compare studies using different methodologies and assessment tools, these discrepancies may depend on specific subcomponents of apathy, which we did not differentiate in this study^3,4,35^.

Our work has limitations. It was a retrospective analysis, albeit drawing on PSP patients in longitudinal observational studies. We relied on clinical diagnosis, although n=64 patients had pathological confirmation of PSP, and clinicopathological confirmation is typically very high in PSP, including in this cohort. Only n=2 patients received a pathological diagnosis that differed from their initial clinical diagnosis of PSP. We also included reports of behavioural features over multiple clinical follow-ups, as it may be that apathy and impulsivity are intermittent rather than constant features. Higher temporal resolution of assessments would be needed to confirm this. Such assessments may rely on carer reports, given a lack of insight of patients into their behavioural changes, which can be present in PSP. There may have been confirmation biases when ascertaining whether patients had certain behavioural features such as apathy and impulsivity. However, we strived to maintain clear selection criteria when assessing these neuropsychiatric problems, drawing on multiple informants and measures.

These used the thresholds set out in Appendix 2 and 3. Another limitation is the use of dichotomic measures of apathy and impulsivity in our study, rather than continuous ratings or variables. This was pragmatic and enabled data to be drawn from a larger cohort, and over a longer period of time. Our cross-sectional findings need replication across other sites and in longitudinal studies and would benefit from systematic simple measures with a dynamic range suitable for patients with PSP. Given the cross-sectional nature of our findings, it is plausible that we may have underestimated the prevalence of apathy and impulsivity. However, these neuropsychiatric features are often present early in PSP^29,36^ and our average follow-up was 5.3 years from symptom onset.

To conclude, our study highlights the co-morbid nature of apathy and impulsivity in PSP and their independence from emotional lability. This informs future research of the neural correlates of apathy and impulsivity, their risk factors, and new therapeutic strategies to reduce apathy and impulsivity.

## Data Availability

data are not available

## Acknowledgement

We thank our patients for their participation in this study. We also thank the brain donors and their family.

## Disclosures

### 2) Financial Disclosures for the previous 12 months

JBR serves as an associate editor to Brain and is a non-remunerated trustee of the Guarantors of Brain, Darwin College and the PSP Association (UK). He also provides consultancy to Asceneuron, Biogen, UCB and has research grants from AZ-Medimmune, Janssen, Lilly as industry partners in the Dementias Platform UK. TR has received honoraria from Biogen, Oxford Biomedica and the National Institute for Health and Clinical Excellence (NICE). The other authors have no disclosures to declare.

## Author Roles

1) Research project: A. Conception, B. Organization, C. Execution;

2) Statistical Analysis: A. Design, B. Execution, C. Review and Critique;

3) Manuscript: A. Writing of the first draft, B. Review and Critique.

Author ZQK: 1A, 1B, 1C, 2A, 2B, 3A

Author AGM and TR: 2C, 3B

Author JBR: 1A, 2C, 3B

Author LP: 1A, 1B, 2A, 2C, 3B

## Ethical compliance statement

This study was conducted in accordance with the 1964 Helsinki declaration. Written informed consent was obtained from all participants. Ethical approval was given by the East of England - Cambridge Central Research Ethics Committee for the “Pick’s Disease and Progressive Supranuclear Palsy Prevalence and Incidence” protocol (12/EE/0475) (October 2015) and by the East of England - Essex Research Ethics Committee for the “Diagnosis and prognosis in Progressive Supranuclear Palsy and Corticobasal Degeneration” protocol (07/Q0102/3) (March 2007). We confirm that we have read the Journal’s position on issues involved in ethical publication and affirm that this work is consistent with those guidelines.

## Supplementary Information

**Supplementary Table 1:** Demographic and clinical data of PSP patients with and without apathy and impulsivity.

| Variables | Apathy and impulsivity (N=101) | Impulsivity Alone (N=15) | Apathy Alone (N=20) | Neither apathy nor impulsivity (N=18) | Overall means | F/x^2 | P-values |
| --- | --- | --- | --- | --- | --- | --- | --- |
| **Demographics** |  |  |  |  |  |  |  |
| **Gender (% female)** | 43.6 | 46.7 | 45 | 50 | 45 (0.5) | - | 0.964 |
| **Age at onset** | 68.1 (7.3) | 67.0(7.4) | 68.0 (7.3) | 70.3 (6.9) | 68.4 (7.22) | 0.65 | 0.585 |
| **Clinical subtypes, severity and motor features** |  |  |  |  |  |  |  |
| **PSP-Richardson's syndrome (% , n)** | 65.22 (75) | 10.43 (12) | 14.78 (17) | 9.57 (11) | 74.68 (115) | - | <0.001 |
| **PSP with CBS features (% , n)** | 60 (9) | 6.67 (1) | 20 (3) | 13.33 (2) | 9.74 (15) | - | 0.001 |
| **PSP with predominant frontal presentation (% , n)** | 92.31 (12) | 7.69 (1) | 0 (0) | 0 (0) | 8.44 (13) | - | 0.001 |
| **Primary gait freezing variant of PSP (% , n)** | 42.86 (3) | 14.29 (1) | 0 (0) | 42.86 (3) | 4.55 (7) | - | 0.047 |
| **PSP-Parkinsonism (% , n)** | 33.33 (1) | 0 (0) | 0 (0) | 66.67 (2) | 1.95 (3) | - | 0.000 |
| **PSP-SL variant (% , n)** | 100 (1) | 0 (0) | 0 (0) | 0 (0) | 0.65 (1) | - | 0.000 |
| **Disease duration (years)** | 5.29 (2.5) | 6.77(4.2) | 4.29 (1.8) | 5.58 (4.1) | 5.34 (2.94) | 2.14 | 0.098 |
| **Disease severity (PSPRS scores)** | 34.92 (13.5) | 40.88 (16.7) | 28.33 (11.8) | 29.63 (13.6) | 33.99 (13.8) | 1.73 | 0.168 |
| **Motor symptoms (UPDRS part III motor scores)** | 33.09 (12.7) | 33.11 (7.7) | 28.79 (13.5) | 28.86 (12.5) | 32.01 (12.3) | 0.63 | 0.601 |
| **Cognitive features** |  |  |  |  |  |  |  |
| **ACE-R on first assessment of behavioural changes** | 79.25 (9.3) | 75.18 (14.3) | 81.80 (11.4) | 77.58 (12.2) | 74.97 (10.6) | 0.91 | 0.439 |
| **FAB on first assessment of behavioural changes** | 11.66(3.3) | 11.80(3.0) | 13.25 (2.3) | 12.00 (1.7) | 11.96 (3.1) | 0.57 | 0.639 |

Means (SDs) are provided. For each of the disease subtypes, percentages of each behavioural subgroup are shown, followed by the number of patients in the corresponding subgroup in brackets i.e. percentage (number of patients). The p values were corrected for multiple comparisons. CBS, corticobasal syndrome; PSP-SL, PSP with speech/language disorders; PSPRS, PSP rating scale; UPDRS, Unified Parkinson's Disease Rating Scale; ACE-R, Addenbrooke's Cognitive Examination scale-Revised; FAB, Frontal Assessment Battery.

### Bayesian ANOVAs

We compared subgroups with ‘both apathy and impulsivity’, ‘only apathy, no impulsivity’, ‘only impulsivity, no apathy’, and ‘neither apathy nor impulsivity’. Bayesian ANOVAs were used to examine differences between clinical variables across the various behavioural subgroups. In this study, the null hypothesis (H_0_) is that there is no difference between the behavioural subgroups for the relevant clinical or disease variable. The alternative hypothesis (H_1_) is that there is a difference between the behavioural subgroups for the relevant disease or clinical variable. Priors assumed equipoise between H_0_ and H_1_. The hypotheses are compared in terms of the Bayes factor, BF. The BF_10_ for the null models was >0.99, indicating that the null model was consistently the best model. The Bayes factors indicated very strong evidence in favour of the null model for differences in ACE-R and FAB (BF_01_>10), but there was inconclusive evidence between models for PSPRS (1/3>BF>3).

## Appendix 1: Qualitative selection criteria for apathy, impulsivity, and emotional lability from clinical letters

### Qualitative selection criteria for apathy

- Apathetic
- Reduced or loss of motivation
- Reduced/Lost interest in new activities
- More socially withdrawn
- Loss of initiative
- “Loss of enjoyment and interest in seeing friends and doing things he used to enjoy”
- Less conversational
- No longer engages in conversations around him/her
- “Frequently spends all day in pyjamas” “Resisting suggestions for additional activity” “Tends to sit all day doing very little” “Wants to stay at home all day” “Spend much of the day in bed”
- “Become rather introverted, engages less in conversations”
- “More mellow personality”
- “Sh/he puts little effort into anything”
- “No longer interested in learning new things”

### Qualitative selection criteria for impulsivity

- “Impulsivity” “impulsive behaviour” (most common)
- Marked disinhibition
- **Social behaviours**
  - Socially or sexually inappropriate behaviour/comments
  - Coarser humour
  - Aggressive behaviour towards family members (physically and/or verbally)
  - More bloody minded and more argumentative +/- more temper outbursts
- **Eating behaviours**
  - Compulsive eating behaviours, cramming behaviour (sometimes leading to dysphagia and choking) +/- decline in table manners
  - Hyperphagia/hyperorality i.e. “Tried to swallow large pieces of food instead of cutting and chewing” “Tendency to overfill his mouth, stuffing his mouth and sucking ++ on beaker”
  - Developed/Increasing sweet tooth
- **Motor behaviours**
  - Unsafe behaviour
  - Motor impulsivity/recklessness and delay intolerant: “Getting up to walk despite instability” i.e. “Impatient and unwilling to wait for people to help out with tasks” “Will often stand up on his own even if he is unstable, leading to frequent falls”
  - Impulsive decision-making i.e., “Making impulsive and irrational decisions that put him at risk”; “Rushes into things or jumps to conclusions and makes decisions without regard for consequences”

### Qualitative selection criteria for emotional lability

- Emotional lability
- Emotional incontinence
- Pseudobulbar affect
- Physical manifestations of distress and crying without corresponding inner emotions
- Emotional volatility i.e. “Can get angry or burst into tears without warning”
- Emotional incongruence - “Unexplained bursting into tears and appearing distressed despite not being internally distressed”, “Tears without feeling sad” “Laughing and crying without feeling happy or sad”
- “Can get tearful or emotional on frequent occasions without any clear triggers to this”
- Laughing/crying out of context
- Inappropriate laughing or crying

## Appendix 2: Cambridge Behavioural Inventory score thresholds for apathy and impulsivity

### CBI Score Thresholds for Apathy

Total score of 1 or more (scoring is based on a Likert scale from 0-never, 1-a few times per month, 2-a few times per week, 3-daily to 4-constantly) on the *Motivation* section of the CBI-R questionnaire in any of the following items: 1) shows less enthusiasm for his or her usual interests; 2) shoes little interest in doing new things; 3) fails to maintain motivation to keep in contact with friends or family; 4) appears indifferent to the worries and concerns of family members; 5) shows reduced affection.

### CBI Score Thresholds for Impulsivity

Total score of 1 or more (scoring is based on a Likert scale from 0-never, 1-a few times per month, 2-a few times per week, 3-daily to 4-constantly) in any of the following items on the *Abnormal Behaviour* section of the CBI-R questionnaire: 1) find humour or laughs at things others do not find funny; 2) has temper outbursts; 3) is uncooperative when asked to do something; 4) shows socially embarrassing behaviour; 5) makes tactless or suggestive remarks; 6) acts impulsively without thinking.

#### And/or

Total score of 1 or more of the following items on the *Eating Habits* section of the CBI-R questionnaire: 1) prefers sweets foods more than before; 2) wants to eat the same food repeatedly; 3) her/his appetite is greater, s/he eats more than before; 4) table manners are declining e.g., stuffing food into mouth

## Appendix 3: PSPRS (Progressive Supranuclear Palsy Rating Scale) score thresholds for apathy, impulsivity, and emotional lability

### Progressive Supranuclear Palsy - Thresholds for apathy, impulsivity, and emotional lability

Total score of 1 or more on the *Withdrawal* section of the PSPRS for apathy, total score of 1 or more on the *Irritability* section of PSP-RS for impulsivity, total score of 1 or more on the *Emotional incontinence* section of the PSPRS for emotional lability.

